# Utilization of Recommended Postnatal Care Services in Baglung District: Barriers, Facilitators, and Implications for Safe Motherhood Programs-A Cross-sectional Study

**DOI:** 10.1101/2025.08.18.25333947

**Authors:** Shijan Acharya, Erina Khatri, Roshani Poudel

## Abstract

According to World Health Organization (WHO), globally 2.4 million children died in the first month of life in 2020. Most neonatal deaths (75%) occur during the first week of life. In 2019, about 1 million newborns died within the first 24 hours. A large proportion of maternal and neonatal deaths occur during the first 48 hours of delivery. Postnatal care (PNC) service utilization plays an important role to reduce maternal and neonatal deaths. In Nepal, the proportion of mothers attending three PNC visits as per the protocol is 25.1 percent in Fiscal Year (FY) 2077/78. In Nepal, Madhesh and Gandaki Province has the lowest PNC use as per protocol in FY 77/78. Several factors hinder the PNC service utilization however the underlying causes remain unclear despite the ongoing efforts of the Government of Nepal (GoN). The major objective of this study was to assess the facilitators and barriers for the uptake of recommended PNC services among women of Baglung district. It is a cross-sectional study. Data collection was done in two randomly selected Rural/Municipalities using multistage cluster sampling technique in January 2024. The findings from this study are useful for safe motherhood program of Gandaki province targeting interventions that are conducive to overcoming PNC utilization barriers. This will ultimately support to increase the PNC utilization as per the protocol of GoN.

## Introduction

The postnatal period, beginning immediately after the birth of the baby and extending up to six weeks (42 days), is a critical time for women, newborns, partners, parents, caregivers and families (1). Reproduction marks the joy of a lifetime for the mother as well as her family. However, this physiological phenomenon is associated with a number of possible outcomes that include mortality of the mother and her baby along with the risk of disability, especially in middle and low-income countries (2).

Globally, there are approximately 6700 newborn deaths daily. The causes of these neonatal deaths were mostly: preterm birth, childbirth-related complications i.e birth asphyxia, infections and birth defects (3). Neonatal deaths and stillbirths arise from poor maternal health, inadequate care during pregnancy, inappropriate management of complications during pregnancy and delivery, poor hygiene during delivery and the first critical hours after birth, and lack of newborn care. Cultural factors such as women’s status in society, their nutritional status at the time of conception, early childbearing, short birth spacing and harmful practices such as inadequate cord care, letting the baby stay wet and cold, discarding colostrum and feeding other foods, also interact in ways that are not always clearly understood.

According to World Health Organization (WHO), more than 70% of all maternal deaths are due to haemorrhage, infection, unsafe abortion, hypertensive disorders of pregnancy, and obstructed labour which are mostly preventable. The major causes of fatalities are poverty; inadequate, inaccessible, and unaffordable health care; low status of women, and illiteracy (4). Inaccessibility to and poor quality of maternal, neonatal and child health (MNCH) services including poor economic condition are among the major factors for the lack of MNCH service use (5).

Maternal deaths pose tremendous adverse psycho-social effects on the families and the quality of life of their children and dependents. This in turn affects the wellbeing of her family and hence, the society(6). Yet, during this period, the burden of maternal and neonatal mortality and morbidity remains unacceptably high, and opportunities to increase maternal well-being and to support nurturing newborn care have not been fully utilized. Postnatal care services are a fundamental component of the continuum of maternal, newborn and child care, and key to achieving the Sustainable Development Goals (SDGs) on reproductive, maternal and child health, including targets to reduce maternal mortality rates and end preventable deaths of newborns (1).

In Nepal, Maternal mortality ratio (MMR) is 151 per 100,000 live births in 2022 whereas neonatal mortality rate is stagnant i.e 21 per 1000 live births since 2016 till 2022 (7). Nepal is facing challenges to decrease neonatal mortality. The percentage of pregnant women attending at least 4 ANC visits as per the protocol was 55.4 percent in Fiscal Year (FY) 2077/78 whereas the proportion of mothers attending three PNC visits as per the protocol was 25.1 percent (lowest in Madhesh and Gandaki Province) in FY 2077/78 which is the lowest among various indicators of safe motherhood program (8). There was a steady increase in Nepal’s proportion of women attending four ANC visits as per protocol, reaching 94% in FY 2079/80. The proportion of institutional deliveries have increased by 13.4% reaching 83.4% in this fiscal year, with 80% deliveries attended by skilled birth attendants and skilled health personnel. Additionally, 44% mothers attended three postnatal checkup (PNC) visits. This gap from ANC to PNC service uptake is being tackled with emphasis on counselling at each point of contact and integration of PNC services with immunization and family planning (9). The proportion of mothers attending three Postnatal Check Up (PNC) visits as per the protocol increased remarkably by nearly 16.0% points from FY 2077/78 to 2078/79. In FY 2079/80, this increased by nearly 3.0% points with 44.0% PNC coverage of three PNC visits. Lumbini (68.0%), Sudurpaschim (61.0%) and Karnali (57.0%) provinces surpassed the national average. Among remaining four provinces, Gandaki Province is one of the provinces with low PNC coverage (9).

All the local units in Baglung have a lower proportion of PNC service utilization compared to the national average (8). Several factors hinder the MNH service utilization, some of which might differ in different parts of the country based on various factors such as socio-economic conditions. Still the reasons for poor PNC utilization remain unclear despite the ongoing efforts of the government and local organizations to promote PNC utilization. The needs and experiences of women in Baglung regarding the MNH service utilization, including PNC services, have not been specifically addressed in the studies published earlier that attempt to identify barriers and facilitators in particular to the PNC utilization in this province. This study attempted to identify the current facilitators and barriers for PNC service utilization of Baglung. This study tried to access the existing factors associated to the PNC service utilizaiton in Baglung and the findings provide the evidences on the gaps.

International safe motherhood initiatives have been applying evidence-based interventions to improve maternal health by increasing accessibility and quality of maternal health services. The WHO advises a minimum of four postnatal care visits. For births in a healthcare facility, both the mother and newborn should receive postnatal care at the facility for at least 24 hours after delivery. If the birth takes place at home, the first postnatal visit should occur as soon as possible within the first 24 hours. Additionally, at least three more postnatal checkups are recommended for healthy mothers and newborns; between 48 and 72 hours, between 7 and 14 days, and around the sixth week after birth. For optimal maternal and newborn health outcomes, healthcare providers should ensure that women and newborns remain in the facility for at least 24 hours following a vaginal birth. Before discharging them, health workers should evaluate key factors, including: a) the physical health of both the mother and baby, as well as the mother’s emotional well-being; b) the mother’s ability to care for herself, along with the confidence and skills of parents and caregivers in looking after the newborn; and c) the home environment and other factors that may impact the mother and baby’s care and access to medical assistance if needed.

It is recommended that skilled health professionals or trained community health workers conduct home visits during the first week after birth for postnatal care. If home visits are not possible or preferred, outpatient postnatal checkups should be arranged (1). Implementation efforts also must include raising awareness of the importance of postnatal care, informing women about what each postnatal care contact entails, and of the fundamental human right of women and newborns to receive postnatal care for their health and well-being. Indicators to monitor progress should capture experience of care and well-being, to enable continuous quality improvement (1,2).

Nepal’s success with reducing maternal mortality is linked to the government’s will regarding maternal health, its promotion of postnatal care, and its focus on community-based delivery of care. Both demand- and supply-side interventions have played important roles in improving maternal and newborn health throughout the country. Policymakers need conceptually and methodologically robust, and generalizable data on how mobile technology might effectively improve the delivery of essential healthcare services for populations in developing nations that are difficult to reach and have limited resources in order to justify public investment (10).

Nepal is lagging behind in reaching the Sustainable Development Goal (SDG) of 90% of women attending three PNCs in accordance with protocol and 70 MMR per 100,000 live births, given the poor rate of PNC utilization relative to ANC visits (11). Likewise, as identified by the Nepal Health Sector Strategy (NHSS) 2015-2020, the gaps in equity and quality of care are the areas to be considered in achieving the SDG 3 (12). Therefore, Government of Nepal (GoN) also has identified MNCH as a priority program and Ministry of Heath and Population (MoHP) has introduced total 4 PNCs including two home visits by health workers to improve PNC utilization (10). Access to prompt PNC services helps to minimize risks of maternal and neonatal mortality and treat any complications arising from the delivery (13).

## Materials and Methods

### Study design, setting, population, and sampling procedure

A descriptive cross-sectional study was conducted to assess the factors associated to PNC utilization among mothers of child < 1year. The study population was women who experienced childbirth during the last 1 year and have completed 42 days of postpartum period. Two rural/municipality of Baglung District was study sites; namely: Baglung Municipality and Tamankhola Rural Municipality (clusters). Data collection was done from total 12 wards (6 from each municipality) from these two (clusters) i.e Tamankhola RM and Baglung M selected randomly.

Multistage cluster sampling was done to select two clusters randomly. The respondents were chosen by lottery method from each unit of the clusters using random sampling.

The required sample size was based on the following formula:

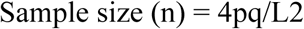

Where P (Percentage of neonates who had 4 PNC check-ups as per protocol in Tamankhola RM (HMIS 2079/80) was 0.237 and Q (Percentage of women who did not have 3 PNC check-ups as per protocol) was 0.736. L (allowable error) was set at 5%.

Sample size required was= 4×0.237×0.736×0.05^2 =279.09 ~ 280 For finitie population,

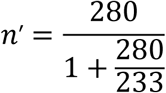

(where, sample size (n)= 280 and expected live birth =233)

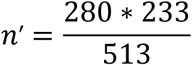

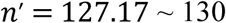

Design effect = 128×1.5= 195

Adding 10 % non response rate final sample size was **215** ∼**219**

### Data collection methods, tool validity, and measurement

Data collection was done through face to face Interview with the mother of under 1 year who were living in selected site for more than 1 year. The tools and guides were translated in Nepali language. Semi-structured questionnaire was prepared based on relevant study (14) was used to assess the knowledge, norms, and experiences relating to PNC service utilization. Reliability of the tool was optimized by translation of English version of the questionnaire (English-Nepali-English-Nepali) by language experts. The questionnaires were translated to Nepali language back and forth and further adapted after pre-testing in the similar context and final questionnaire was consulted with expert working in Maternal and Neonatal Health (MNH) section of Family Welfare Division (FWD).

### Ethical consideration

This study included human participation hence ethical approval of this study was obtained from Institutional Review Committee (IRC) of Institute of Medicine (IOM). Approvals from the health office, the respective rural/municipality and participants was taken before data collection and objective of the study was clarified. The written informed consent signed by participants who are willing to take part in the study was taken prior to data collection. In case of pregnant women less than 18 years, assent form was signed from participant and consent was taken from the participant’s guardian. Obtained data was coded and analyzed to maintain anonymity of respondents. The research title is sensitive in itself as the inquiry related to health of mother and child; their experiences were asked as respondents might not feel comfortable to answer. The respondents who had experienced complicated delivery and/or loss of life. This was dealt sensitively by the researcher. Female researcher conduced data collection and they was oriented before data collection in order to be sensible while asking questions without making the participants uncomfortable.

### Statistical analysis

Responses were recoreded in mobile application Commcare and exported to Ms Excel. Futher after data cleaning it was exported to IBM SPSS Statistics version 23 for analysis and coding. For categorical variables: frequencies, percentages was done. For continuous variables: mean and standard deviation or median and inter-quartile range. Chi-square test or to find the association between independent variables PNC utilization as per protocol. During analysis, at 95% confidence interval, a p-value of less than 0.05 was considered to be significant. Binary logistic regression was done for p<0.1 for Multivariate analysis.

## Results

A total of 219 data were collected to assess the status on PNC utilization from women having an infant child (<12 months) and have completed post partum period of 42 days.

### 4.1.1 Socio-demographic characteristics of the study participants

Of the total 219 study participants, the median age of women were 25years with IQR 8. The majority (178, 81.3%) of the respondents were in the age group of 20-34 years. Majority 23.7% of women had education of lower secondary education. Most of the respondents (63.9%) were Dalit and (210, 95.9%) of the respondent follow hindu religion. Among 219 respondents almost 39% respondend that they live in nuclear family. The majority (93, 42.5%) of the respondent were multipara having two children. Majority of respondents (45.2%) were homemaker and as caretaker of their children. Regarding the occupation of husband majority (79, 36.1%) had foreign employment in countries like Japan, India. The major source of income of the family was found to be agriculture (61.1%) The median monthly income was Nepalese Rupees (NPR) 30,000 and Inter Quartile Range (IQR) was NPR 20,000

**Table 1.**
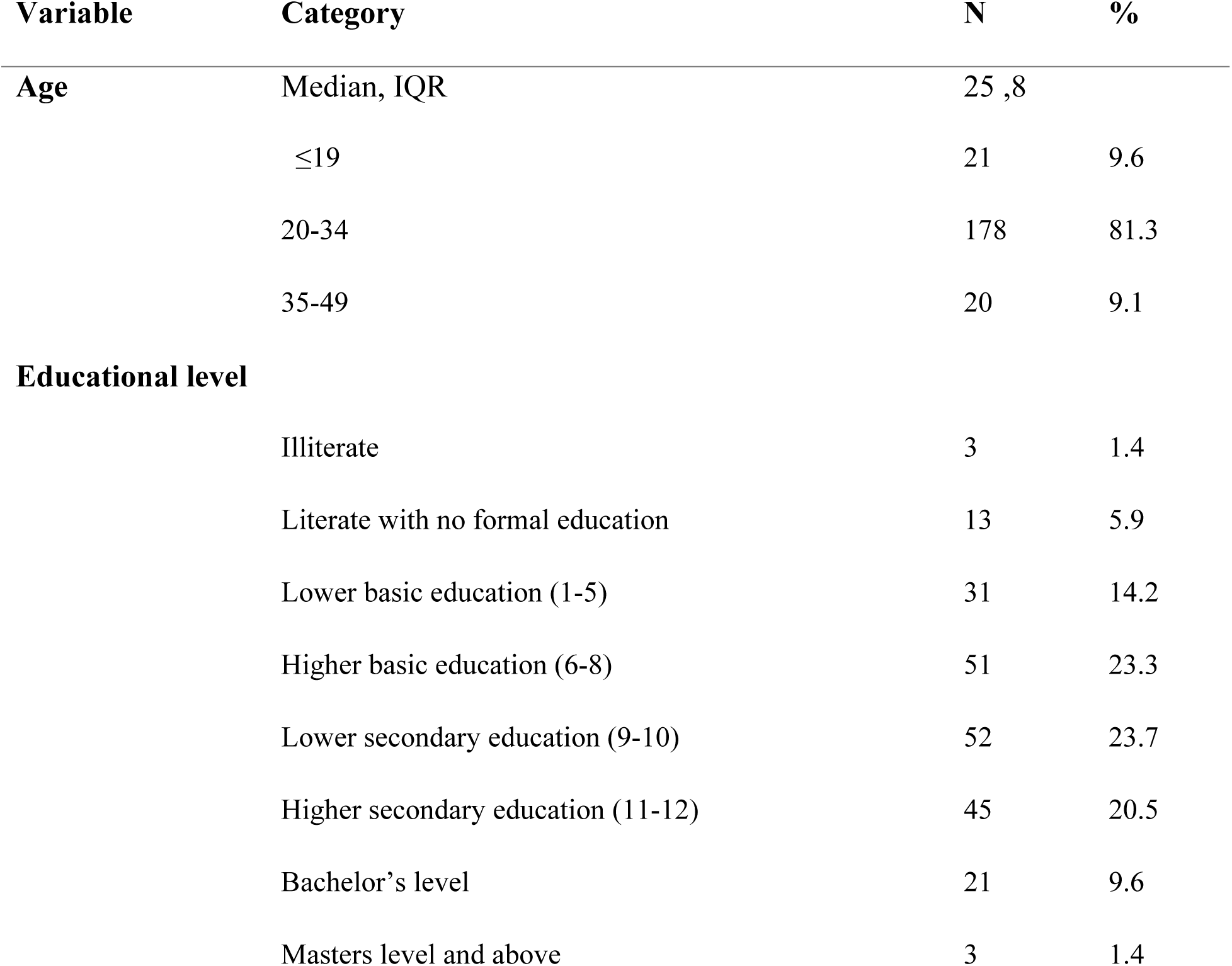
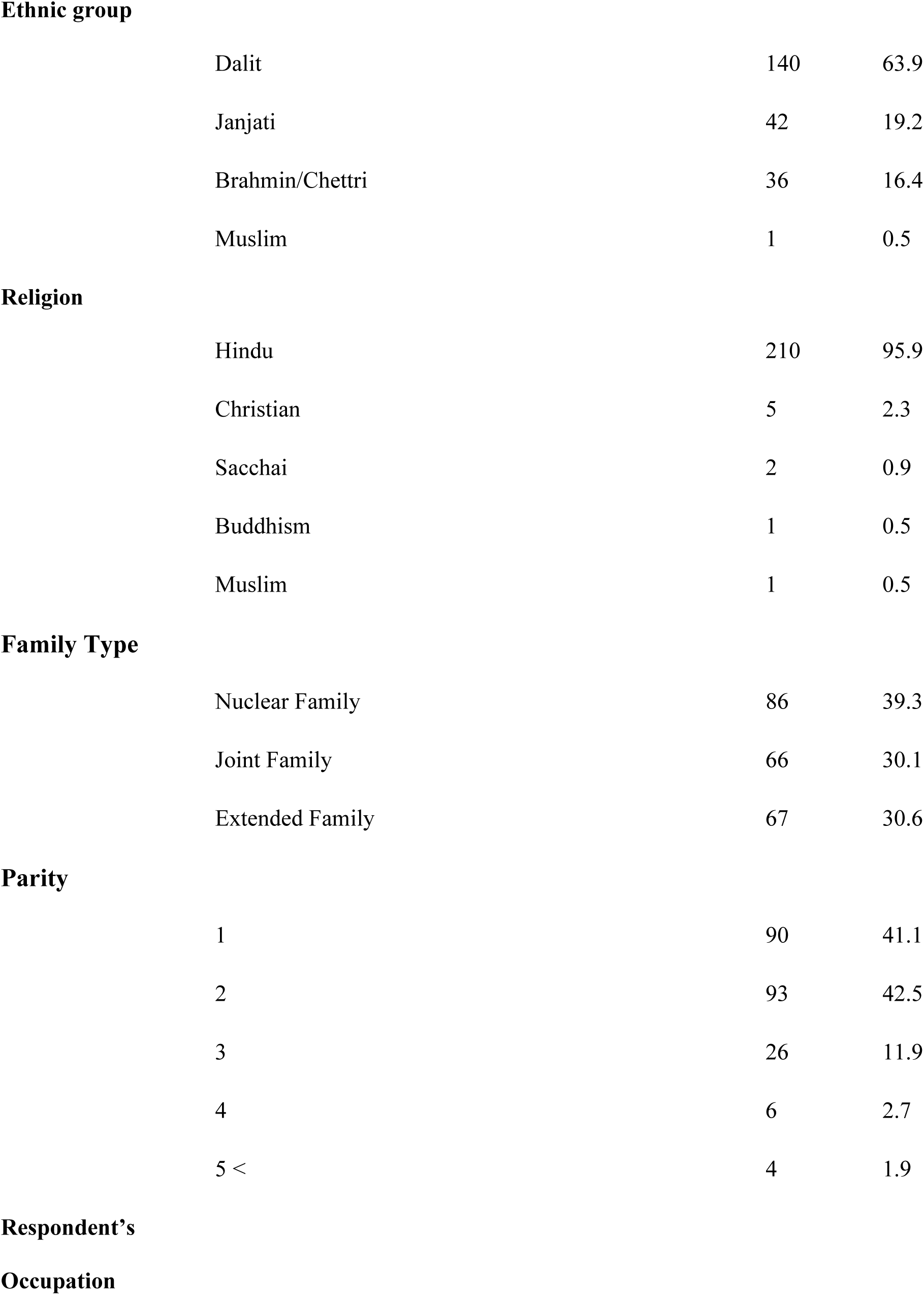
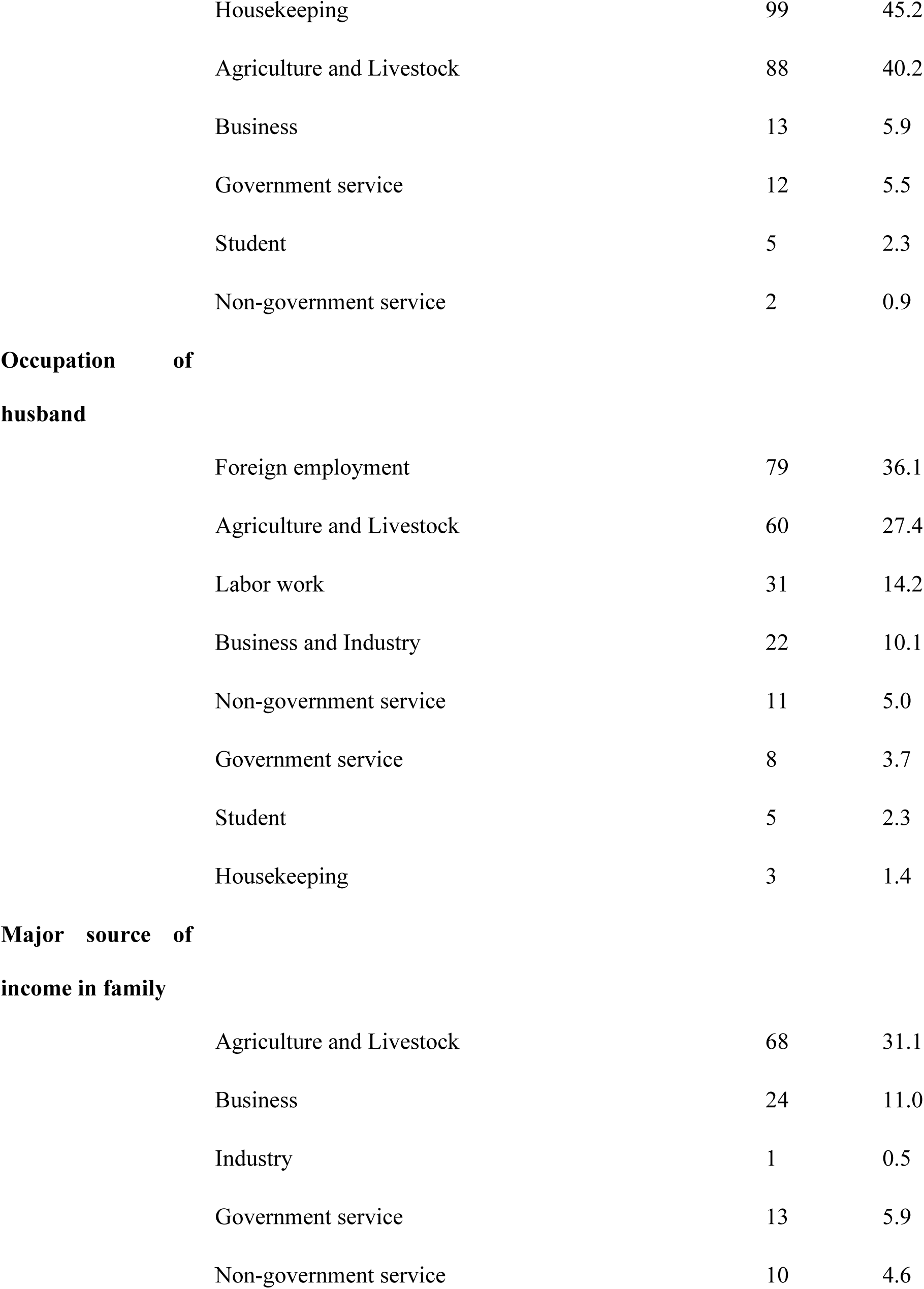
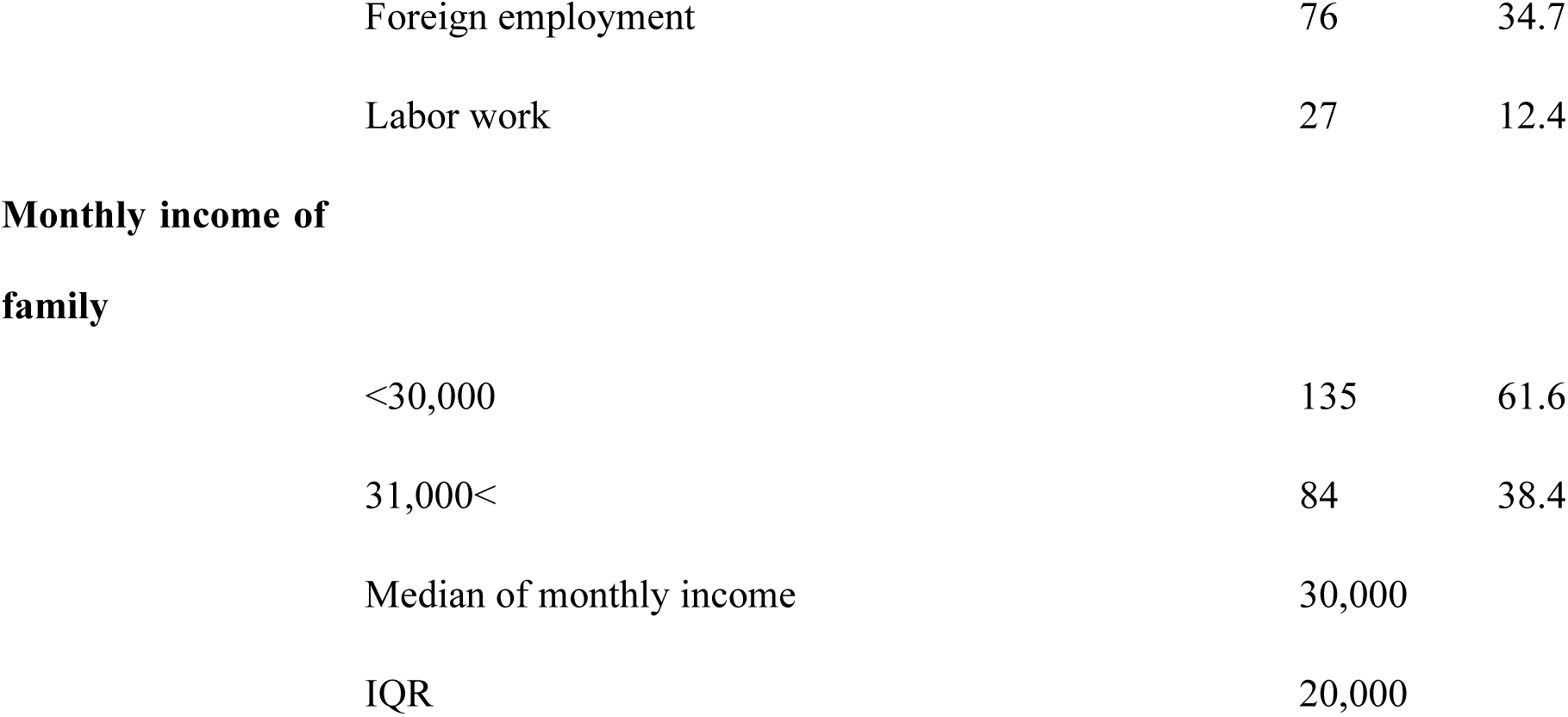
Sociodemographic characteristics of the study participants (n=219)

| Variable | Category | N | % |
| --- | --- | --- | --- |
| Age | Median, IQR | 25 ,8 |  |
|  | ≤19 | 21 | 9.6 |
|  | 20-34 | 178 | 81.3 |
|  | 35-49 | 20 | 9.1 |
| **Educational level** |  |  |  |
|  | Illiterate | 3 | 1.4 |
|  | Literate with no formal education | 13 | 5.9 |
|  | Lower basic education (1-5) | 31 | 14.2 |
|  | Higher basic education (6-8) | 51 | 23.3 |
|  | Lower secondary education (9-10) | 52 | 23.7 |
|  | Higher secondary education (11-12) | 45 | 20.5 |
|  | Bachelor's level | 21 | 9.6 |
|  | Masters level and above | 3 | 1.4 |

**Ethnic group**
|  |  |  |
| --- | --- | --- |
| Dalit | 140 | 63.9 |
| Janjati | 42 | 19.2 |
| Brahmin/Chettri | 36 | 16.4 |
| Muslim | 1 | 0.5 |

**Religion**
|  |  |  |
| --- | --- | --- |
| Hindu | 210 | 95.9 |
| Christian | 5 | 2.3 |
| Sacchai | 2 | 0.9 |
| Buddhism | 1 | 0.5 |
| Muslim | 1 | 0.5 |

**Family Type**
|  |  |  |
| --- | --- | --- |
| Nuclear Family | 86 | 39.3 |
| Joint Family | 66 | 30.1 |
| Extended Family | 67 | 30.6 |

**Parity**
|  |  |  |
| --- | --- | --- |
| 1 | 90 | 41.1 |
| 2 | 93 | 42.5 |
| 3 | 26 | 11.9 |
| 4 | 6 | 2.7 |
| 5 < | 4 | 1.9 |
| **Respondent's****Occupation** |  |  |  |
|  | Housekeeping | 99 | 45.2 |
|  | Agriculture and Livestock | 88 | 40.2 |
|  | Business | 13 | 5.9 |
|  | Government service | 12 | 5.5 |
|  | Student | 5 | 2.3 |
|  | Non-government service | 2 | 0.9 |
| **Occupation of husband** |  |  |  |
|  | Foreign employment | 79 | 36.1 |
|  | Agriculture and Livestock | 60 | 27.4 |
|  | Labor work | 31 | 14.2 |
|  | Business and Industry | 22 | 10.1 |
|  | Non-government service | 11 | 5.0 |
|  | Government service | 8 | 3.7 |
|  | Student | 5 | 2.3 |
|  | Housekeeping | 3 | 1.4 |
| **Major source of income in family** |  |  |  |
|  | Agriculture and Livestock | 68 | 31.1 |
|  | Business | 24 | 11.0 |
|  | Industry | 1 | 0.5 |
|  | Government service | 13 | 5.9 |
|  | Non-government service | 10 | 4.6 |

|  |  |  |
| --- | --- | --- |
| Foreign employment | 76 | 34.7 |
| Labor work | 27 | 12.4 |
| **Monthly income of family** |  |  |
| <30,000 | 135 | 61.6 |
| 31,000< | 84 | 38.4 |
| Median of monthly income | 30,000 |  |
| IQR | 20,000 |  |

This table clearly presents the data with categories and sub-categories under each variable of the demographic and socioeconomic characteristics of the study participants. Here is an analysis of the frequencies, highlighting the highest percentages within each category. The majority of respondents (81.3%) are in the 20-34 age group.

This indicates that the majority of respondents are relatively young adults. The highest percentage is for Lower secondary education (9–10), with 23.7% of respondents falling into this category. Higher basic education (6–8) also accounts for a significant portion at 23.3%. This shows that most respondents have a secondary education or equivalent. The highest percentage is for the Dalit ethnic group, with 63.9% of respondents identifying as such. This group forms the largest segment of the sample, suggesting it is a dominant ethnic group in this population. The highest percentage is for Hindu respondents, who make up 95.9% of the sample. The highest percentage is for Nuclear Family, with 39.3% of respondents living in this family structure. This is closely followed by Extended Family (30.6%) and Joint Family (30.1%). The highest percentage is for those with 2 children, accounting for 42.5% of respondents. This suggests that having two children is the most common family size among the respondents. The next most common family size is 1 child, with 41.1% of respondents. The highest percentage is for Housekeeping, with 45.2% of respondents identifying this as their occupation. This suggests a predominant role for women or homemakers in the sample population. Agriculture and Livestock is also significant, with 40.2% of respondents working in this sector. The highest percentage is for foreign employment, which accounts for 36.1% of respondents’ husbands. This indicates that many of the respondents’ husbands are employed abroad, which might suggest a migration trend in the study area. The second largest category is Agriculture and Livestock, with 27.4%. The highest percentage comes from Foreign employment, with 34.7% of respondents indicating it as their primary source of income. This further supports the prominence of foreign migration as a significant economic factor. Agriculture and Livestock also plays a key role, with 31.1% of respondents identifying it as the primary income source. The highest percentage is for families earning <30,000, which make up 61.6% of the sample. This suggests that a majority of families in the study live on relatively low monthly incomes.

### MNH service utilization status

Of the total 219 respondents almost 47% still resided more than half an hour to three hours away from basic health services. Regarding the ANC use 99% had visited health facility at least once during their pregnancy period, whereas 72% had done ANC visits as per the protocol (i.e more than eight times as recommended) during their pregnancy period. Regarding the delivery 217 (99%) of the respondents had perform their delivery at health institution. After the child birth among the 219 mothers from home delivery and institutional delivery only 52 of them had done PNC checkup as per the government protocol (i.e at least four times)

**Table 2.**
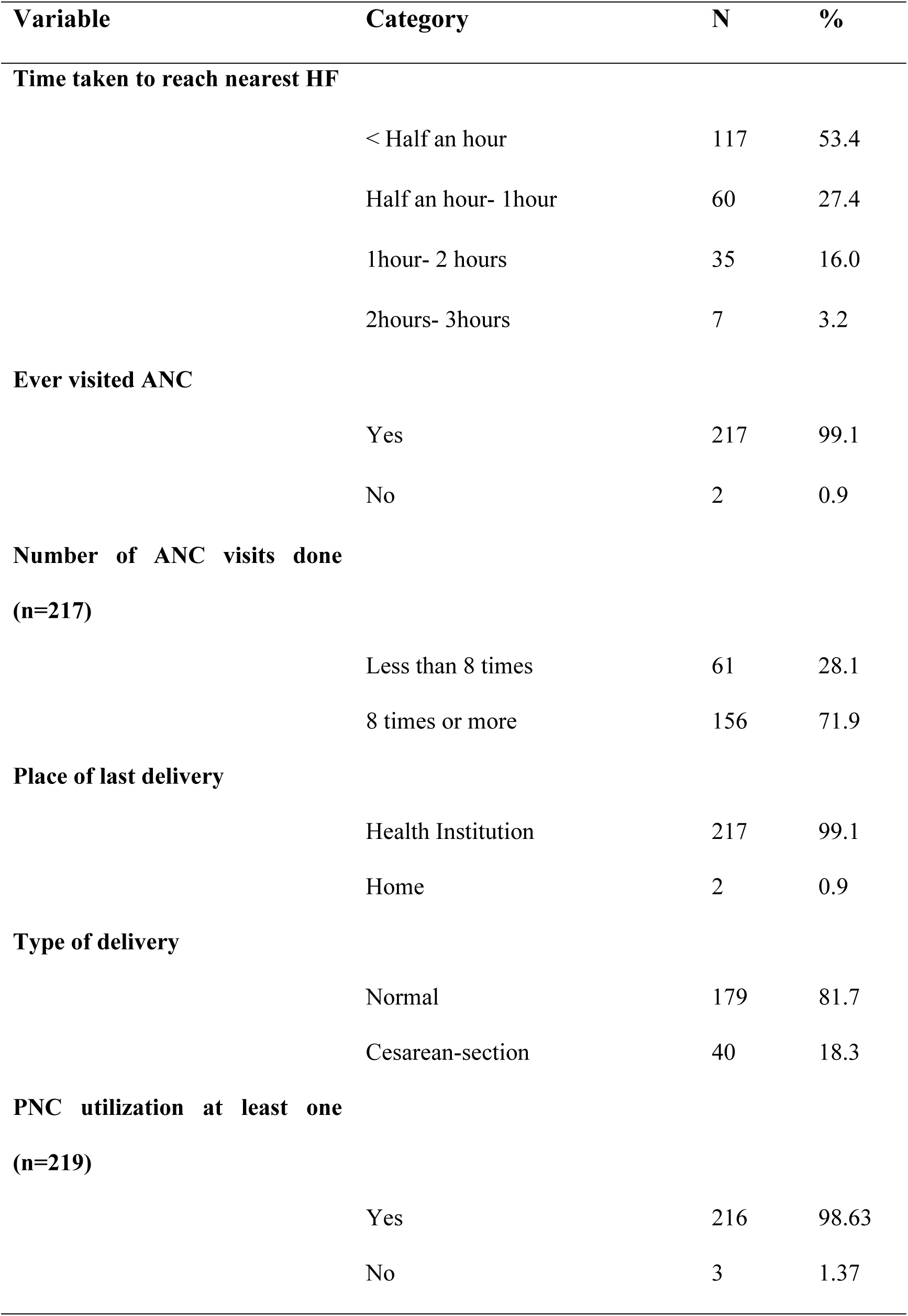
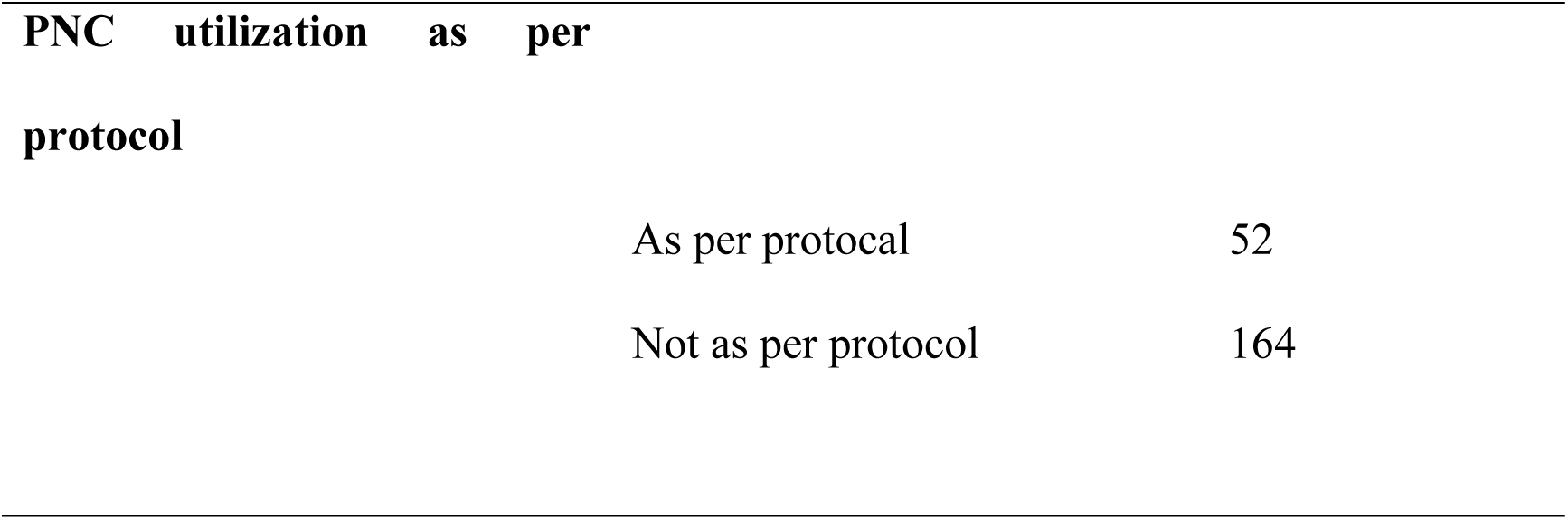
ANC and PNC service utilization.

| Variable | Category | N | % |
| --- | --- | --- | --- |
| **Time taken to reach nearest HF** |  |  |  |
|  | < Half an hour | 117 | 53.4 |
|  | Half an hour- 1hour | 60 | 27.4 |
|  | 1hour- 2 hours | 35 | 16.0 |
|  | 2hours- 3hours | 7 | 3.2 |
| **Ever visited ANC** |  |  |  |
|  | Yes | 217 | 99.1 |
|  | No | 2 | 0.9 |
| **Number of ANC visits done**<br>**(n=217)** |  |  |  |
|  | Less than 8 times | 61 | 28.1 |
|  | 8 times or more | 156 | 71.9 |
| **Place of last delivery** |  |  |  |
|  | Health Institution | 217 | 99.1 |
|  | Home | 2 | 0.9 |
| **Type of delivery** |  |  |  |
|  | Normal | 179 | 81.7 |
|  | Cesarean-section | 40 | 18.3 |
| **PNC utilization at least one**<br>**(n=219)** |  |  |  |
|  | Yes | 216 | 98.63 |
|  | No | 3 | 1.37 |

**PNC utilization as per protocol**
|  |  |
| --- | --- |
| As per protocol | 52 |
| Not as per protocol | 164 |

### Knowledge regarding Postnatal Care

Regarding the knowledge on postnatal checkup, among the total 219 respondents 210 (96 %) have heard about the Postnatal care and among those 210 respondents only 16.7 % have responded that Post natal check up should be done at least four times after the child birth. Of the total respondent 94 % have knowledge regarding the danger sign for mothers during the post natal period and around 96% have knowledge on danger sign for baby during the postnatal period.

**Table 3.**
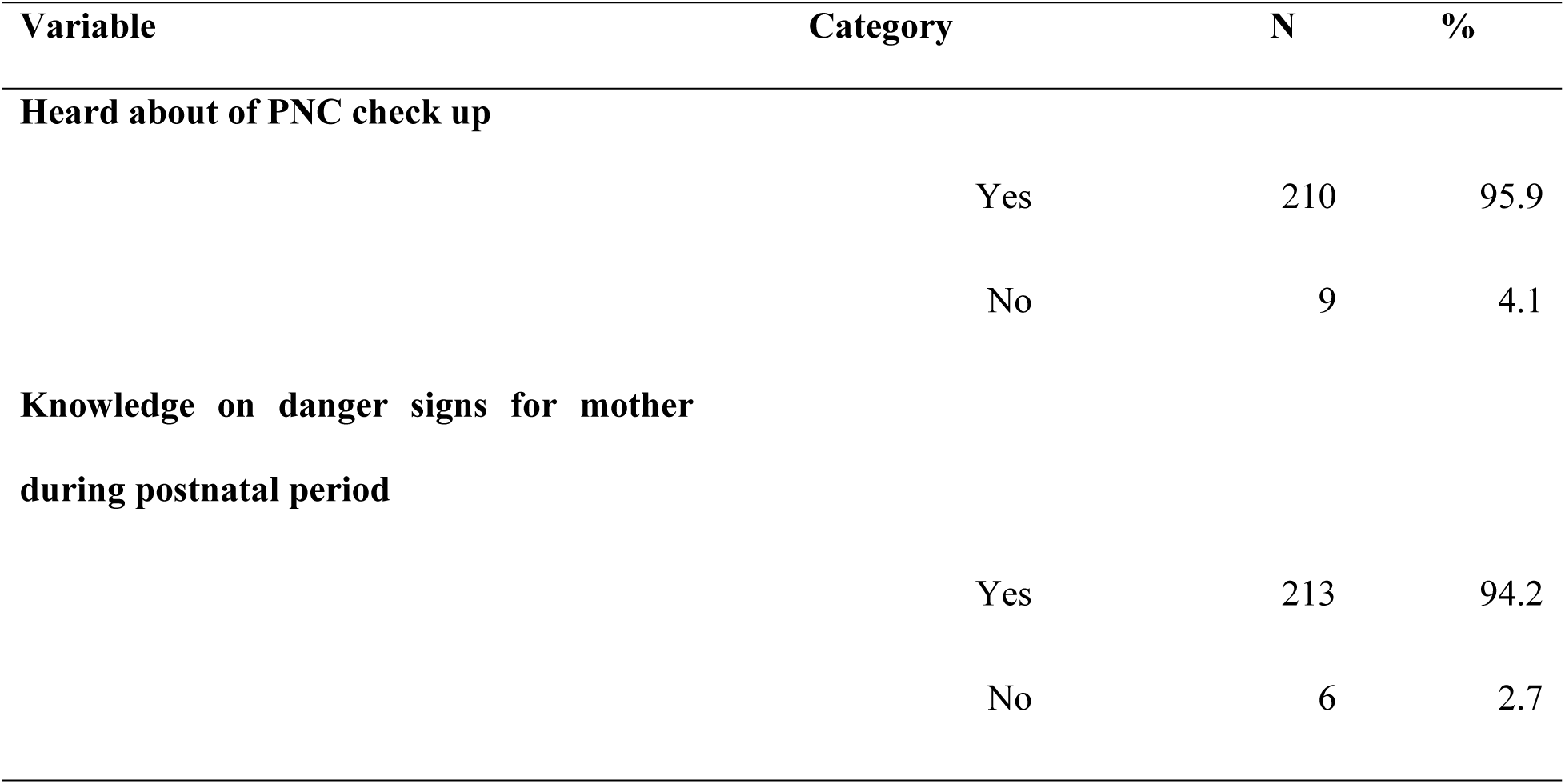
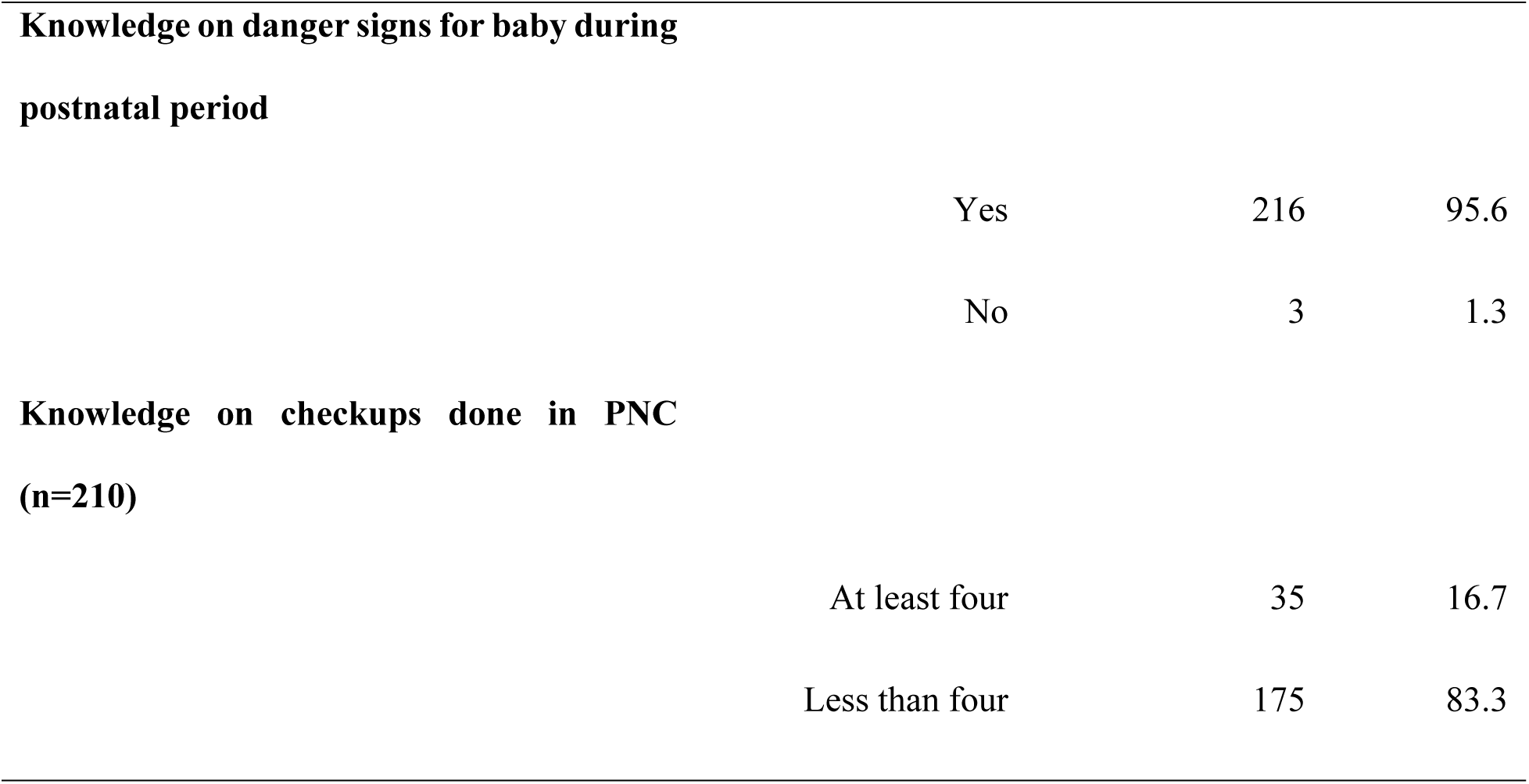
Knowledge regarding Postnatal Care.

| Variable | Category | N | % |
| --- | --- | --- | --- |
| **Heard about of PNC check up** |  |  |  |
|  | Yes | 210 | 95.9 |
|  | No | 9 | 4.1 |
| **Knowledge on danger signs for mother**<br>**during postnatal period** |  |  |  |
|  | Yes | 213 | 94.2 |
|  | No | 6 | 2.7 |

**Knowledge on danger signs for baby during postnatal period**
|  |  |  |
| --- | --- | --- |
| Yes | 216 | 95.6 |
| No | 3 | 1.3 |

**Knowledge on checkups done in PNC (n=210)**
|  |  |  |
| --- | --- | --- |
| At least four | 35 | 16.7 |
| Less than four | 175 | 83.3 |

In order to assess the feasibility of Government intervention on Pregnancy and PNC tracking and follow-up through SMS few questions were asked based on the use of mobile phone to assess the readiness for such intervention in future. Regarding the use of mobile phone, when asked if they owned a mobile phone, 96% responded that they had mobile phone whereas 96% used it for receiving call followed by 87% of women who could read SMS in Nepali language.

**Table 4.** Use of mobile phone.

| **Characteristics** | **Frequency** | **Percentage** |
| --- | --- | --- |
| Own a mobile phone |  |  |
| **Yes** | 211 | 96.3 |
| **No** | 8 | 3.7 |
| Uses of mobile phone (n=211) |  |  |
| **Receiving call** | 210 | 95.9 |
| **Reads SMS** | 194 | 88.6 |

### Association between socio-demographic characteristics with PNC utilization

For the bivariate analysis, chi-square test was performed between independent and dependent variable. The age, educational level, ethnic group, religion was compared in relation to PNC utilization as per protocol, where none of the characteristics were found to be significant. Bivariate analysis was done for the variables such as: occupation of women, occupation of husband, major source of income and monthly income of the family compared in relation to PNC utilization as per protocol, where none of the characteristics were found to be significant. Family type, parity, time taken to reach the nearest HF, ANC utilization was also found to be not significant.

**Table 5.** Bivariate analysis on socio-demographic characteristics and PNC utilization.

| Variables | PNC utilization n (%) |  | Crude OR<br>(C.I.) | p-value |
| --- | --- | --- | --- | --- |
|  | Yes | No |  |  |
| Age (n=219) |  |  |  |  |
| ≤19 | 4 (19.0) | 17 (81) | Ref |  |
| ≥20 | 48 (24.2) | 150 (75.8) | 1.4 (0.44-4.24) | 0.596 |
| Educational level |  |  |  |  |
| No formal education | 6 (37.5) | 10 (62.5) | Ref |  |
| Formal education | 46 (22.7) | 157 (77.3) | 0.5 (0.17-1.42) | 0.187 |
| Ethnic group |  |  |  |  |
| Dalit | 34 (24.3) | 106 (75.7) | Ref |  |
| Non-Dalit | 18 (22.8) | 61 (77.2) | 0.9 (0.48-1.77) | 0.802 |
| Occupation of women (n=219) |  |  |  |  |
| Not earning | 21 (20.2) | 83 (79.8) | Ref |  |
| Earning | 31 (27) | 84 (73) | 1.5 (0.77-2.74) | 0.241 |
| **Occupation of husband** |  |  |  |  |
| Foreign employment | 19 (24.4) | 59 (75.6) | Ref |  |
| In-country employment | 33 (23.4) | 108 (76.6) | 0.9 (0.49-1.81) | 0.874 |
| **Monthly income of family** |  |  |  |  |
| <30,000 | 34 (25.2) | 101 (74.8) | Ref |  |
| 31,000< | 18 (21.4) | 66 (78.6) | 0.8 (0.42-1.55) | 0.526 |
| **Parity** |  |  |  |  |
| Primi para | 16 (17.8) | 74 (82.2) | Ref |  |
| Multi para | 36 (27.9) | 93 (72.1) | 1.8 (0.922-3.475) | 0.085 |

### Association between socio-demographic characteristics with PNC utilization

The bivariate analysis is done for the variables such as: type of delivery, place of delivery compared in relation to PNC utilization as per protocol, where none of the characteristics were found to be significant. The bivariate analysis is done between access mobile phone compared in relation to PNC utilization as per protocol where the significance was seen among the mobile users and PNC utilization, which means that those who had phone were 0.2 times less likely to utilize PNC service as per protocol as compared to those who did not own mobile phone. This relation of significance might have been influenced by the number of women who did not had mobile phone were only 8 in number, among which most of them resided nearby health facilities or were in contact with either health worker or FCHV, which influenced their behavior in utilization of PNC services.

**Table 6.**
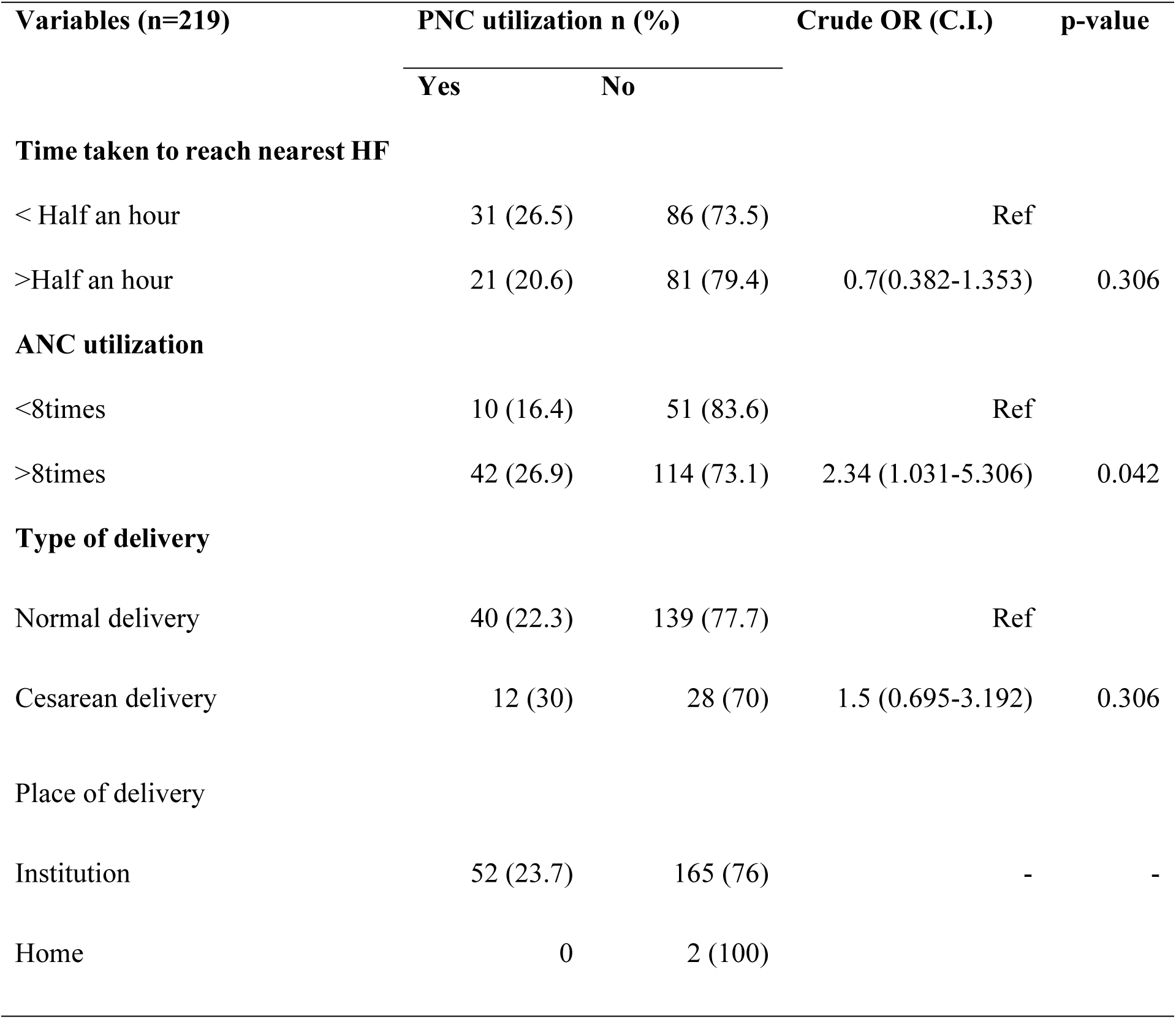
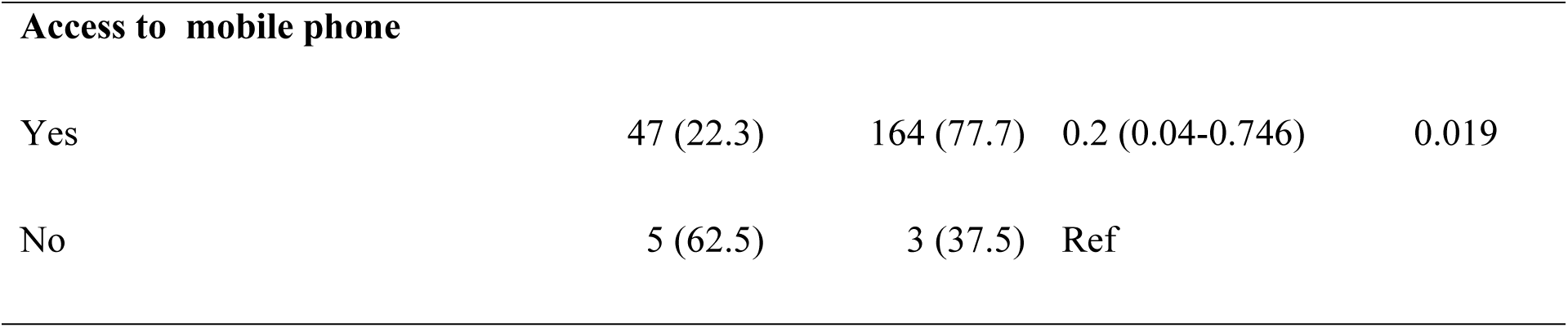
Bivariate analysis on MNC service utilization and PNC utilization.

Besides, further study must be done to explore what kind of information does women seek in mobile phone or internet, or those who are educated mostly use mobile phone for various purposes, and might opt for advanced services this might influence the low rate of PNC utilization. However, further study needs to be done to explore the reasons behind it.

The bivariate analysis is done for the variables such as: type of delivery, place of delivery compared in relation to PNC utilization as per protocol, where none of the characteristics were found to be significant. The bivariate analysis is done between having mobile phone compared in relation to PNC utilization as per protocol. Bivariate analysis of mobile phone use and PNC service utilization. The significance was seen among the mobile users and PNC utilization, which means that those who had phone were 0.2 times more likely to utilize PNC service as per protocol as compared to those who did not own mobile phone. This relation of significance might have been influenced by the number of women who did not had mobile phone were only 8 in number, among which most of them resided nearby health facilities or were in contact with either health worker or FCHV, which influenced their behavior in utilization of PNC services.

Besides, further study must be done to explore what kind of information does women seek in mobile phone or internet, or those who are educated mostly use mobile phone for various purposes, and might opt for advanced services this might influence the low rate of PNC utilization. However, further study needs to be done to explore the reasons behind it.

Similarly, In the given Table 20, the bivariate analysis is done for the variables such as: identified risks for mother, baby, regarding knowledge of PNC and advise given during ANC visit.

**Table 7.** Bivariate analysis on knowledge, risk assessment and PNC service utilization.

| Variables | PNC utilization n (%) |  | Crude OR<br>(C.I.) | p-value |
| --- | --- | --- | --- | --- |
|  | Yes | No |  |  |
| Risk assessment during PNC check for child |  |  |  |  |
| Risk identified and referred | 4 (17.4) | 19 (82.6) | - | - |
| No risk identified | 48 (25) | 144 (75) |  |  |
| Risk assessment during PNC check for mother |  |  |  |  |
| Risk identified and referred | 6 (35.3) | 11 (64.7) | - | - |
| No risk identified | 46 (23.2) | 152 (76.8) |  |  |
| Heard about danger signs during PNC for mother |  |  |  |  |
| Yes | 50 (23.5) | 163(76.5) | Ref |  |
| No | 2 (33.3) | 4 (66.7) | 0.6 (0.11-3.45) | 0.579 |
| Heard of PNC services |  |  |  |  |
| Yes | 50 (23.8) | 160 (76.2) | Ref |  |
| No | 2 (22.2) | 7 (77.8) | 1.1 (0.22-5.44) | 0.913 |
| Advised for PNC utilization during ANC visit |  |  |  |  |
| Yes | 50 (25) | 150 (75) | - | - |
| No | 2 (11.1) | 16 (88.9) |  |  |

In the given Table 20, the bivariate analysis done for the variables such as: identified risks for mother, baby, regarding knowledge of PNC and advise given during ANC visit, where none of the characteristics were found to be significant.

After the bivariate analysis the p-value less than 0.3 were taken for multivariate analysis where having phone and ANC utilization variables were seen significant. As for ANC utilization, as shown in given Table 21, those women who had utilized ANC more than eight times as per protocol recommended by the GoN, PNC utilization as per protocol among them were found to utilize the PNC services 2.34 (aOR: 2.34 (1.03-5.30), <0.05) times more than those utilized ANC service less than 8 times. Similarly, those who owned mobile phone were 0.14 (aOR: 0.14 (0.30-0.64), <0.05) times more likely to utilize PNC services as compared to those who did own mobile phone.

**Table 8.** Multivariate analysis on mobile phone use, ANC visit with respect to PNC utilization.

| Characteristics | PNC utilization n (%) |  | aOR (C.I.) | p-value |
| --- | --- | --- | --- | --- |
|  | Yes | No |  |  |
| Own mobile phone |  |  |  |  |
| Yes | 47 (22.3) | 164 (77.7) | 0.14 (0.30-0.649) | 0.012 |
| No | 5 (62.5) | 3 (37.5) | Ref |  |
| ANC utilization |  |  |  |  |
| <8times | 10 (16.4) | 51 (83.6) | Ref |  |
| >8times | 42 (26.9) | 114 (73.1) | 2.34 (1.031-5.306) | 0.042 |

The Hosmer-Lemeshow Chi-square statistic in the model showed no significant difference (p=0.756) between observed and expected probabilities test. Hence, the goodness-of-fit of this model was not poor. The Negelkerke R^2^ value was noted to be 0.103 which indicates that 10 percent of variability in the PNC utilization as per protocol was explained by the independent variables.

**Table 9.** Bivariate analysis on knowledge, risk assessment and PNC service utilization.

| Characteristics | PNC utilization |  | Crude OR<br>(C.I.) | p-value |
| --- | --- | --- | --- | --- |
|  | n (%) |  |  |  |
|  | Yes | No |  |  |
| Risk assessment during PNC check for child |  |  |  |  |
| Risk identified and referred | 4 (17.4) | 19 (82.6) | - | - |
| No risk identified | 48 (25) | 144 (75) |  |  |
| Risk assessment during PNC check for mother |  |  |  |  |
| Risk identified and referred | 6 (35.3) | 11 (64.7) | - | - |
| No risk identified | 46 (23.2) | 152 (76.8) |  |  |
| Heard about danger signs during PNC for mother |  |  |  |  |
| Yes | 50 (23.5) | 163(76.5) | Ref |  |
| No | 2 (33.3) | 4 (66.7) | 0.6 (0.11-3.45) | 0.579 |
| Heard of PNCservices |  |  |  |  |
| Yes | 50 (23.8) | 160 (76.2) | Ref |  |
| No | 2 (22.2) | 7 (77.8) | 1.1 (0.22-5.44) | 0.913 |

| Advised for PNC utilization during ANC visit |  |  |  |  |
| --- | --- | --- | --- | --- |
| Yes | 50 (25) | 150 (75) | - | - |
| No | 2 (11.1) | 16 (88.9) |  |  |

In the given Table 20, the bivariate analysis done for the variables such as: identified risks for mother, baby, regarding knowledge of PNC and advise given during ANC visit, where none of the characteristics were found to be significant.

After the bivariate analysis the p-value less than 0.3 were taken for multivariate analysis where having phone and ANC utilization variables were seen significant. As for ANC utilization, ashown in given Table 21, those women who had utilized ANC more than eight times as per protocol recommended by the GoN, PNC utilization as per protocol among them were seen high and was significantly proven.

**Table 10.**
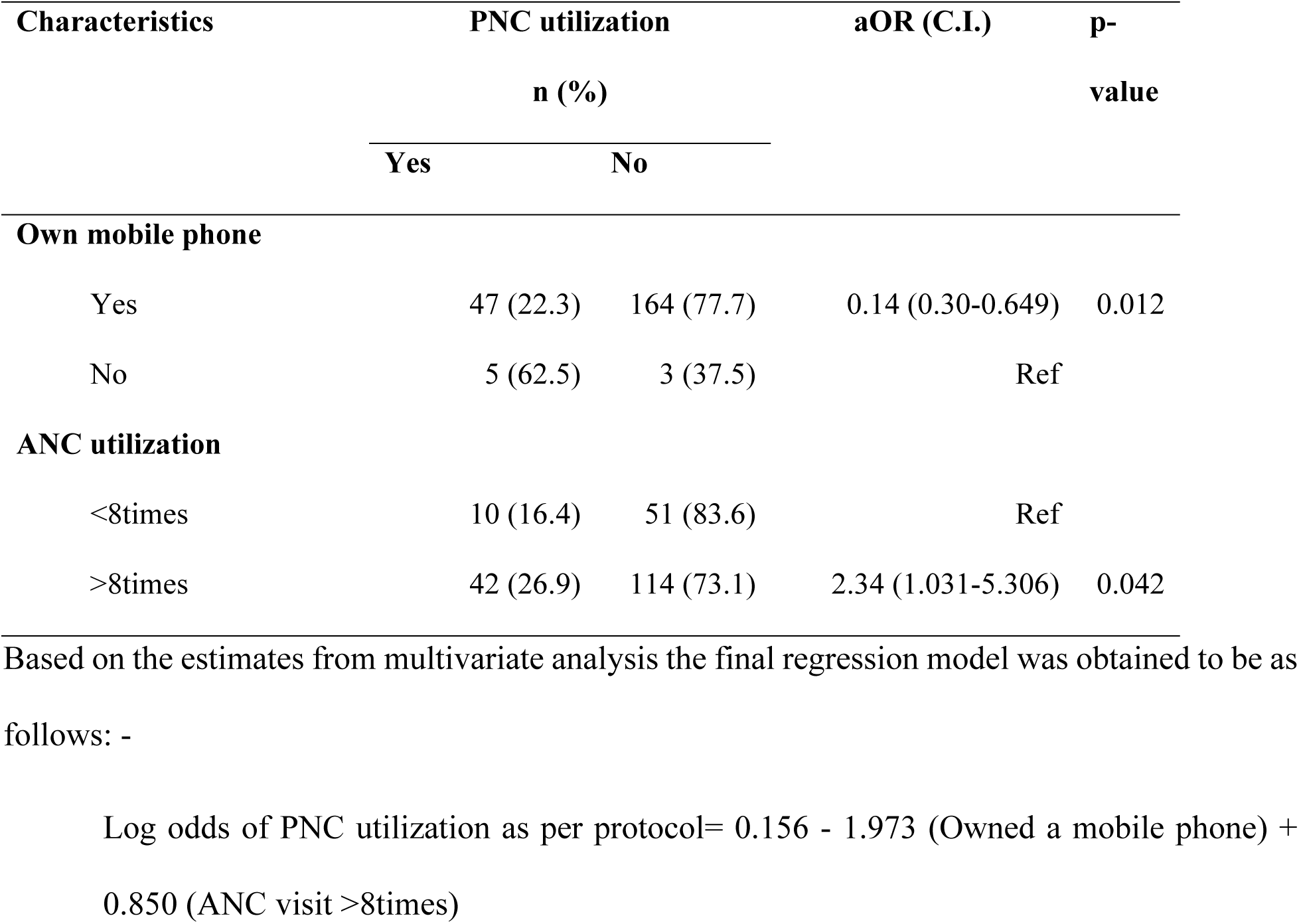
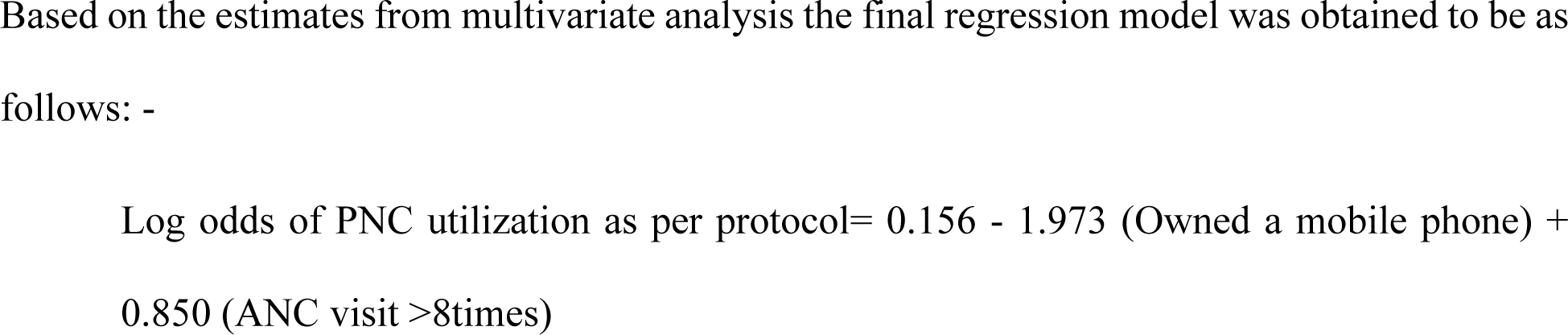
Multivariate analysis on mobile phone use, ANC visit with respect to PNC utilization.

The Hosmer-Lemeshow Chi-square statistic in the model showed no significant difference (p=0.756) between observed and expected probabilities test. Hence, the goodness-of-fit of this model was not poor. The Negelkerke R^2^ value was noted to be 0.103 which indicates that 10 percent of variability in the PNC utilization as per protocol was explained by the independent variables.

The safemotherhood program of Nepal is focusing on 100% institutional delivery whereas, according to the finding of this study, 1% still had home delivery. Both the home delivery was in Tamankhola RM where intervention was carried out. Almost 99% had utilized at least one PNC, as 99% had institutional delivery therefore they received their first PNC at health facility. However, after the delivery the facility stay varied from one hour to few days depending on the type of delivery. It was found that women wished to return to their home after delivery and the health workers did not make it mandatory for the health facility stay for at least 24 hours.

The first PNC as per protocol was 100% among those who had done at least one PNC followed by the second PNC which was 70.4% followed by third PNC which was 67.1%, followed by 4^th^ PNC which was 34.7 %. The second and third PNCs should be done by Nursing staffs by visiting home was 58% in second visit whereas it was 80% in third visit as mostly CS cases had to stay longer in health facility, while sometime health workers and FCHVs were not informed regarding their return, if they had opted to deliver the baby in elsewhere.

Total 96% had heard about the PNC check ups among which only 17% could tell at least four checkups or services given to mother or child under PNC services. Among which most of them know about the immunization service. Total 94% knew the danger signs of mother during the postnatal period, among them only 25% could tell at least four danger signs for mother during postnatal period, most of them could tell about post partum haemmorhage. Total 96% knew the danger signs of baby during postnatal period.

When asked about the knowledge on the available PNC services for both the mother and newborn most of them ie 35% knew about immunization service provided followed by growth monitoring of child i.e 10%, followed by counseling services regarding dos and don’ts during postnatal period i.e 9.7%. Very few i.e 2.4% knew about the family planning services and the application of navi malam. Total 15% responded other PNC services that were not listed for example: advanced health services in case of complication like severe sickness of newborn after delivery or other health conditions.

As per the PNC guideline by GoN, women should be able to tell at least four danger signs of mother among the listed danger signs. Out of total 25% could tell post partum haemorrhage as the danger sign followed by high temperature or fever 14.7%, followed by vaginal tearing and bleeding (11.2%), breast abscess (10%), very few recognized anaemias as the danger signs during post partum period.

Regarding the danger signs of newborn, 23% could tell temperature or fever followed by difficulty in respiration (17%), irregularity in breastfeeding (44%), whereas only few could mention about Jaundice (5.4%) and change in colour, movement and irritable crying of baby (5.3%) as danger signs.

Regarding the use of mobile phone, when asked if they owned a mobile phone, 96% responded that they had mobile phone whereas 96% used it for receiving call followed by 87% who could read SMS in Nepali language however most of them responded they do not get any kind of messages from anywhere usually.

## Discussion

PNC services are important and have been found to be an influential indicator of MNH use (15)(16)(17). Among MNH services, PNC services are the least utilized. In this study, PNC as per protocol was found to be 23.7% identical to the HMIS data of Baglung district (18). In this study only 36% women could decide by herself to utilize MNCH services both for herself and her child. Majorly decision making was jointly done by spouse followed by sole decisions made by in-laws and husbands similar to the various studies. Yet, as previous researches have shown, a small percentage of women claimed to have decided to seek healthcare services on their own (15).

The primary source of income was foreign employment, which is why husbands held financial positions and had more decision-making authority followed by mother-in-law (16). This was not, however, considered to be an obstacle for the utlizaiton of ANC services, as the ANC utilization as per protocol was close to 72% (19). The distance to travel to health facility within half an hour were 53% while remaining had to travel more. Home visit interventions aimed at providing accessible postnatal care services to women have been observed to increase PNC utilization; in fact, home visits accounted for 57% of all visits during the second visit and 80% of all visits during the third PNC visit. In order to increase PNC service utilization, a targeted outreach facility might be more successful concentrating on the fourth PNC visit, as it is the lowest of all (19). Total 41% of the participants in a study identified as primi para, which is consistent with previous research indicated that, despite family encouragement, primi para women may choose not to seek care out of shyness, particularly if they are young or become pregnant at an early age, alike to the women from Pakistan and Uganda (19)(20)(17). Seclusion practices have been associated with a delay in seeking PNC service in one study carried out in Nepal; however, in this study cultural barriers were not identified as a barrier (21). Fear of pain was the major reason for feeling susceptibile during childbirth (22).Individuals with more understanding and awareness regarding dangers during perpuerium period planned on using PNC services (19). Similar to this study, in a study done in Ethiopia, majority of respondendts knew danger signs of postnatal period among which the postpartum hemorrhage (bleeding) was the frequently mentioned danger sign. Similar to this study among the mothers who visited health facility within 42 days were for immunization purpose similar to the study done in Ethiopia, where among women who attended PNC, more than two-third, attended mainly to immunize their baby. In this study, the majority, of the respondents mentioned lack of information as a main reason for not utilizing postnatal services similar to the study done in Ethiopia (23). Conclusions and recommendations

## Conclusions

This study mainly focuses on the factors associated to the utilization of PNC services. Notably, credible endorsements from reputable sources, addressing knowledge, such as danger signs during PNC and practice of doing ANC visits encourage for the uptake of recommended PNC. The utilization of PNC services in this study was influenced by several factors. While a majority of women (99%) accessed at least one PNC due to the high institutional delivery rate, adherence to the recommended four PNC visits was significantly lower, with only 34.7% completing all four. Knowledge gaps regarding PNC services and postnatal danger signs were evident, as only a small proportion of women could identify key maternal and newborn complications which suggests limited health education or insufficient counseling during ANC visits.

Socioeconomic factors, including education, family type, and ethnicity, did not show a significant association with PNC utilization. However, women with higher ANC adherence (eight or more visits) demonstrated better compliance with PNC guidelines. Interestingly, mobile phone ownership, while prevalent, was inversely associated with PNC utilization. This unexpected finding may reflect the influence of alternative support systems (e.g., proximity to health facilities or community health worker involvement) among non-users.

Efforts to improve PNC utilization should focus on strengthening health education, especially regarding danger signs and the importance of follow-up visits, during antenatal and delivery services. Targeted interventions for rural or under-informed populations regarding availability of MNH services and recommended PNC could enhance adherence to PNC protocols.

## Data Availability

All relevant data are within the manuscript.

## Acknowledgement

I would like to express my heartfelt gratitude to all those who have provided guidance, support, and encouragement throughout this study. First and foremost, I am deeply thankful to the Central Department of Public Health (CDPH), Institute of Medicine (IOM), Tribhuvan University (TU). My sincere appreciation goes to my supervisor, Assistant Professor Mr. Ashok Bhurtyal, and co-supervisor, Teaching Assistant Ms. Roshani Poudel, for their invaluable supervision and continuous support. I am also grateful to the Head of CDPH, Mr. Amod Poudyal, and all the faculty members for their guidance.

I extend my thanks to the Health Office Baglung for funding this study and providing me with this opportunity. Special thanks to Mr. Baburam Acharya (Head of Health Office Baglung), Mr. Prabin Sharma (former Head of Health Office Baglung), Ms. Shima Kunwar (Public Health Nurse), Ms. Laxmi Sharma (Public Health Inspector), and all other staff of the Health Office Baglung. I also appreciate the support from Mr. Tilak Channtyal and Mr. Ram Prasad Khanal (Health Section Chiefs), as well as the health facility in-charges, nursing staff, technical and non-technical staff, and FCHVs in Tamankhola and Baglung municipalities.

Additionally, I am grateful to the Ministry of Health, Gandaki Province, for creating research opportunities. I would like to express my sincere thanks to all the study participants for their time and cooperation during data collection. Lastly, I extend my gratitude to everyone who contributed, either directly or indirectly, to the success of this study.

## Acronyms

ANC: Antenatal Care
CS: Cesarean Section
FWD: Family Welfare Division
FY: Fiscal Year
GoN: Government of Nepal
HBM: Health Belief Model
MMR: Maternal Mortality Ratio
mhealth: Mobile Health
MoHP: Ministry of Health and Populaton
MPDSR: Maternal and Perinatal Death Surveillance and Response
NDHS: Nepal Demographic Health Survey
NHSS: Nepal Health Sector Strategy
PNC: Postnatal Care
RM: Rural Municipality
SBA: Skilled Birth Attendant
SDG: Sustainable Development Goals
WHO: World Health Organization

## Author Contributions (CRediT Taxonomy)

- **SA**: Conceptualization; Methodology; Investigation; Writing Original Draft; Project Administration
- **EK**: Data analysis; Review and Editing
- **RP**: Formal Analysis; Supervision; Writing, Review & Editin

